# Analysis of COVID-19 Infection Amongst Healthcare Workers in Rivers State, Nigeria

**DOI:** 10.1101/2022.01.31.22270058

**Authors:** CN Eze-Emiri, FA Patrick, EO Igwe, GC Owhonda

**Affiliations:** Department of Epidemiology, School of Public Health, University of Port Harcourt, Rivers State, Nigeria; Department of Public Health & Disease Control, Rivers State Ministry of Health, Port Harcourt, Rivers State, Nigeria; Faculty of Medicine and Health Sciences, University of Wollongong NSW, Australia

**Keywords:** Coronavirus, COVID-19, Healthcare workers, nosocomial infection, severity, mortality

## Abstract

**Background/aims:** Healthcare workers (HCWs) are at an increased risk of infection and mortality associated with the COVID-19 pandemic. This study determined the illness severity and mortality amongst COVID-19 infected healthcare workers.

**Methods:** The current study was a retrospective cohort study using population-level data. Secondary analysis was conducted on collated data from the Public Health Emergency Operations Centre (PHEOC) at the State Ministry of Health. The cohort included all documented healthcare workers with confirmed COVID-19 infection (diagnosed by Polymerase Chain Reaction). Data were gathered from the COVID-19 patient database of the PHEOC, on demographics, place of work, illness severity and outcome. Descriptive statistics were reported on the cohort characteristics. Adjusted odds ratio was used to report the measure of association between illness severity and risk factors.

**Results:** The mean age was 43 years and 50.5% of the cohort were female. Of the 301 healthcare workers, 187 patients were symptomatic with 32 requiring hospitalisation. From the available data, seven infected HCWs died of their COVID-19 infection, resulting in a case fatality ratio of 2.3%. A subgroup analysis was conducted on the health professionals infected –doctors (71.7%), nurses (27.3%), others (1%). Symptomatic cases were more inclined to progress to severe illness. Predictors of mortality assessed included age, sex, case class and illness severity. The logistic regression model was statistically significant, *χ*^2^(9) = 16.965, *α* = 0.049.

**Conclusion:** Frontline healthcare workers are at an increased risk of exposure to COVID-19 infections. In Nigeria, there is a higher risk of experiencing a severe disease if symptomatic while infected with COVID-19. It is imperative that preventive strategies, proper education, and awareness are put in place to protect healthcare workers.

**Summary Box:** Healthcare workers as first responders, are vulnerable to workplace infections. It is manifest in the COVID-19 pandemic where deaths of healthcare workers resulted in further shortage of the already compromised human resource; consequently compromising effective healthcare delivery. As the pandemic progresses, studies have been conducted globally on this topic and scientific evidence continues to show higher mortality and disease severity of COVID-19. Nevertheless, it is important to understand the effect of COVID-19 on healthcare workers in Nigeria-a developing country.

This study highlights the illness severity and mortality associated with COVID-19 among the study population; its results presented a higher case fatality rate than both national and subnational rates. The results also further emphasises the need to protect healthcare workers; ensure they are knowledgable in both infection prevention and control, and that the healthcare space is safe against nosocomial infections

The study adds to the scientific evidence on the severity and mortality associated with COVID-19 in Nigeria. A national research is needed to extrapolate the findings from this study to the nation. Hence, expatiate on the global fight against coronaviruses such as COVID-19.

## INTRODUCTION

Healthcare workers (HCWs) have a higher risk of encountering infectious agents due to their work environments. With the COVID-19 pandemic, frontline HCWs face a higher risk of infection and mortality as well as being drivers of community-level infection. Recent evidence shows that compared to individuals in the general community, frontline HCWs have a 12-fold risk of testing positive for COVID-19, with higher risk observed for workers with inadequate access to personal protective equipment (PPE) ^1^. In addition to increased exposure to COVID-19 in the pandemic, Wang et al. (2020)^2^ found that poor sleep quality and higher working pressure can increase the risk of nosocomial SARS-CoV-2 infection amongst HCWs. Hence, it is possible to extrapolate these results to the Nigerian healthcare setting as the density of HCWs (1.95 to 1,000 persons) in the country is reportedly “still very low” to effectively deliver essential health services ^3^. The estimated mortality rate amongst HCWs attributed to COVID-19 has increased progressively ^4^. In May 2020, the total number of reported HCW deaths from 67 countries was 1413. Consequently, it suggests that for every 100 HCWs that got infected, one died –the deaths were also 0.5% of the total number of 270,426 COVID-19 deaths worldwide ^5^. Additionally, a survey of 37 countries estimated median death of 0.05 HCWs per 100,000 population. A report from the Pan American Health Organization (PAHO) in collaboration with the World Health Organization (WHO) stated that approximately 570,000 healthcare workers got infected with COVID-19 with above 2,500 dying from the virus across the American region alone ^6^. The World Health Organization also estimated that between 80,000 and 180,000 healthcare workers died of COVID-19 in the period between January 2020 to May 2021, converging to a medium scenario of 115,500 deaths ^7^.

A subnational study highlighting the burden of COVID-19 amongst healthcare workers is paramount to understanding the effect of the pandemic on the healthcare workforce in Nigeria. The study aim was to determine the illness severity and mortality amongst COVID-19 infected healthcare workers in Rivers State, Nigeria.

## METHOD

This study was conducted according to the guidelines of the Declaration of Helsinki. The Ethics Committee of the Rivers State Ministry of Health gave approval for this work –Ethics ID: MH/PRS/391/VOL.2/809.

### Study location

The study was conducted in Rivers State, located in the South-South geopolitical zone of Nigeria.

### Study design and population

The study was a retrospective cohort study using population-level data. The cohort included all documented healthcare workers with confirmed COVID-19 infection. The healthcare workers were categorised using the World Health Organization and International Labour Organisation (ILO) International Standard Classification of Occupations (ISCO) ^8^. There were five categories based on their roles in patient management and healthcare services:

- Health professionals –medical doctors, nurses, dentist, pharmacists, health safety professionals.
- Health associate professional –all technologists and assistants in health professions, Community Health Workers.
- Personal care workers –health care and home-based care workers.
- Health management and support personnel –administrative and management staff, trade workers, social workers, life science professionals.
- Other health service providers –armed forces staff, interns, and hospital volunteers.

Furthermore, the health facilities were classified on the basis of services rendered. Hospital classification by services: Teaching hospitals –offering tertiary health services; General hospitals –offering secondary health services; Community hospitals –offering primary and community-based care; specialised outpatient clinics – rendering specialty outpatient services like dentistry, radiology, and diagnostic services; Corporate/Occupational health clinics –offering general and occupational health services, restricted to employees only; and Health allied organisations.

### Data Source

Secondary data was collated from the data reported to the data centre of the Public Health Emergency Operations Centre (PHEOC) at the State Ministry of Health. The data sources included reports from public and private-owned health facilities, containment centres, offshore platforms, and other health-allied facilities. The duration of data extraction was from 24 March 2020 to 30 November 2021. The dataset characterised demographics, pre-existing comorbidities, symptoms, facility managed, patient status, treatment outcome, and dates of related events, without personal identifiers. Hence, this secondary analysis waived the required individual informed consent. Patient information was retrieved from the dataset based on the occupation of interest –healthcare worker and their respective designation—; alongside demographic data on age, sex; other information collected included the place of work defined as ‘health facility’, illness severity defined as ‘hospitalisation required’, case classification defined as ‘symptomatic or asymptomatic’, knowledge of exposure, place of exposure, and treatment outcome labeled as ‘recovered’, ‘alive’, or ‘dead’.

### Data analysis

Data were analysed using IBM SPSS Statistics for Windows, Version 25 ^9^. Descriptive statistics were used to report on the cohort characteristics. Means and standard deviations were reported for continuous variables and proportions for categorical and qualitative response variables. Univariate analysis of categorical variables was conducted using Chi-square (*χ*^2^) –and Fischer’s exact test where appropriate. A two-tailed p-value less than 0.05 was statistically significant. Multivariate logistic regression was used to evaluate risk factors of illness severity and mortality among healthcare workers with COVID-19. The adjusted odds ratio (aOR) with 95% confidence interval (95% CI) was used to report the measure of association between the following: illness severity and risk factors –age, sex, and case class; mortality and risk factors –age, sex, illness severity and case class.

## RESULTS

### Patient Characteristics

Data on 301 healthcare workers infected with COVID-19 were identified and extracted to a spreadsheet. Demographic and clinical characteristics of the patients is found in Table 1. The mean age was 43 years and 50.5% of the cohort were female. Of the 301 healthcare workers, 187 patients were symptomatic with 10% (32) of the study cohort requiring hospitalisation-a measure of illness severity. 108 healthcare workers were in contact with known probable cases, and 101 persons knowing the place of exposure. From the available data, 7 (26.4%) infected HCWs died of their COVID-19 infection, resulting in a case fatality ratio of 2.3%. Table 2 shows a subgroup analysis conducted on the health professionals infected –doctors (71.7%), nurses (27.3%), others (1%); and teaching hospitals by ownership: public (78.6%), private (21.4%). The distribution of healthcare workers by the World Health Organization classification^8^ and health facilities is located in figure 1 and 2, respectively.

**Table 1.** Demographics and characteristics of COVID-19 amongst healthcare workers (n = 301)

| **VARIABLE** | **n (%)** |
| --- | --- |
| **SEX** |  |
| Male | 149 (49.5) |
| Female | 152 (50.5) |
| **AGE** | 43 ± 11.7* |
| 20–29 | 25 (8.3) |
| 30–39 | 118 (39.2) |
| 40–49 | 81 (26.9) |
| 50–59 | 44 (14.6) |
| 60–69 | 23 (7.6) |
| >70 | 10 (3.3) |
| **CASE CLASS** |  |
| Symptomatic | 187 (62.1) |
| Asymptomatic | 114 (37.9) |
| **ILLNESS SEVERITY** (requiring hospitalization) |  |
| Yes | 32 (10.6) |
| No | 269 (89.4) |
| **CONTACT WITH PROBABLE CASE** |  |
| Yes | 108 (35.9) |
| No | 156 (51.8) |
| Non-response/Incomplete data | 37 (12.3) |
| **KNOWLEDGE OF SUSPECTED EXPOSURE** |  |
| Yes | 101 (33.6) |
| No | 157 (52.3) |
| Non-response/Incomplete data | 47 (15.7) |
| **EXPOSURE** |  |
| Church | 12 (4) |
| Home | 12 (4) |
| Social event | 26 (8.7) |
| Workplace | 51 (17) |
| **OUTCOME** |  |
| Recovered | 294 (97.7) |
| Dead | 7 (2.3) |

\*Mean ± S.D, Case fatality ratio, CFR = 2.33%

**Table 2.** Subgroup analysis.

|  |  |
| --- | --- |
| **Health professionals** | **240 (100)** |
| Doctors | 172 (71.7) |
| Nurses | 63 (26.3) |
| **Hospitals by ownership** | **276 (100)** |
| Public | 217 (78.6) |
| Private | 59 (21.4) |

**Table 3.** Outcome proportion by risk factors.

| Variables, n (%) | Illness severity (n=32) | mortality (n=7) |
| --- | --- | --- |
| **Age** |  |  |
| 20–29 | 2 (6.3) | 0 (0) |
| 30–39 | 9 (28.1) | 1 (14.3) |
| 40–49 | 13 (40.6) | 0 (0) |
| 50–59 | 4 (12.5) | 2 (28.6) |
| 60–69 | 3 (9.4) | 3 (42.9) |
| >70 | 1 (3.1) | 1 (14.3) |
| **Sex** |  |  |
| Male | 16 (50) | 6 (85.7) |
| Female | 16 (50) | 1 (14.3) |
| **Case class** |  |  |
| Symptomatic | 30 (93.8) | 5 (71.4) |
| Asymptomatic | 2 (6.3) | 2 (28.6) |
| **Illness severity** |  |  |
| Yes | - | 1 (14.3) |
| No | - | 6 (85.7) |

**Figure 1.**
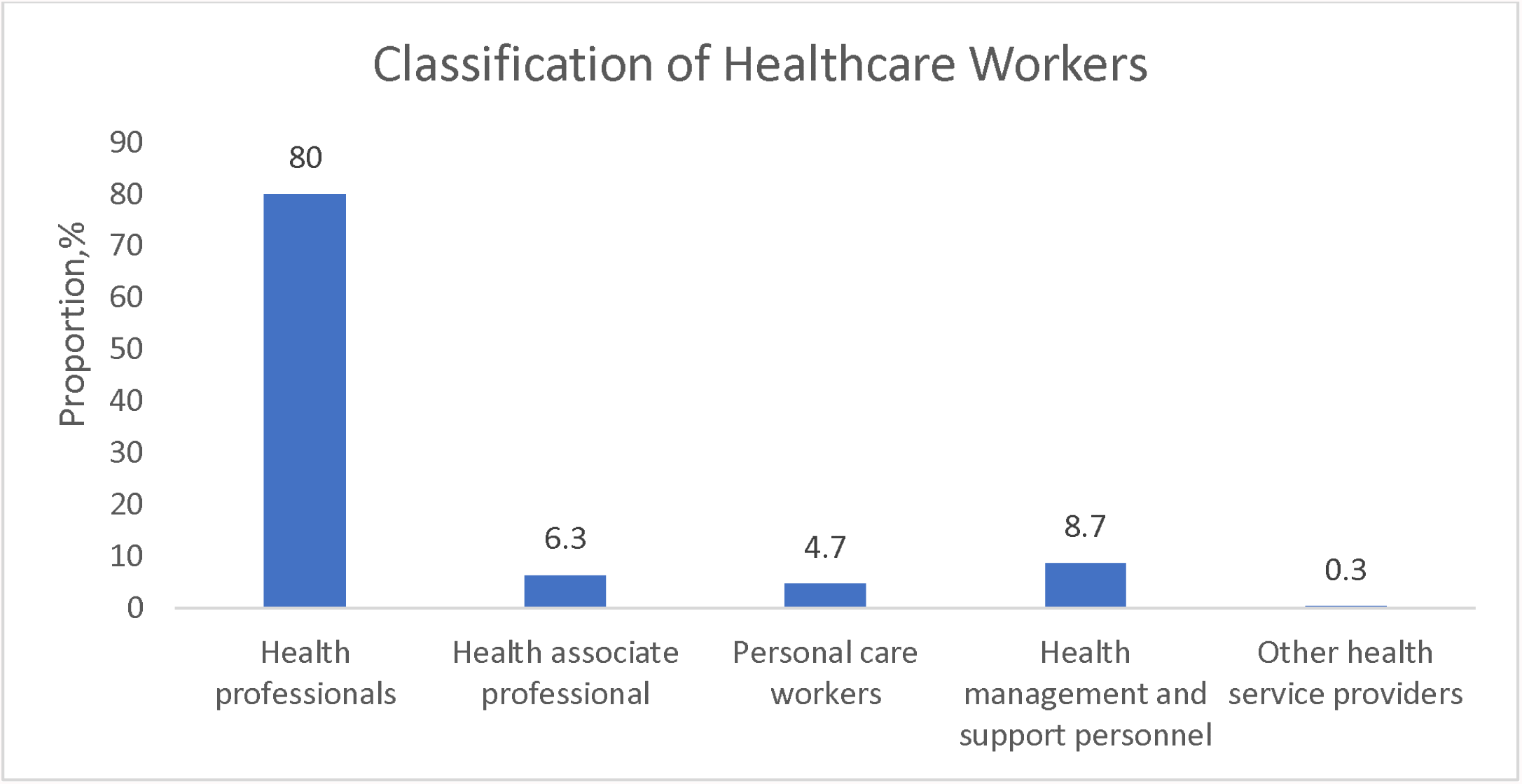
The distribution of infected Healthcare workers by classification^8^.

**Figure 2.**
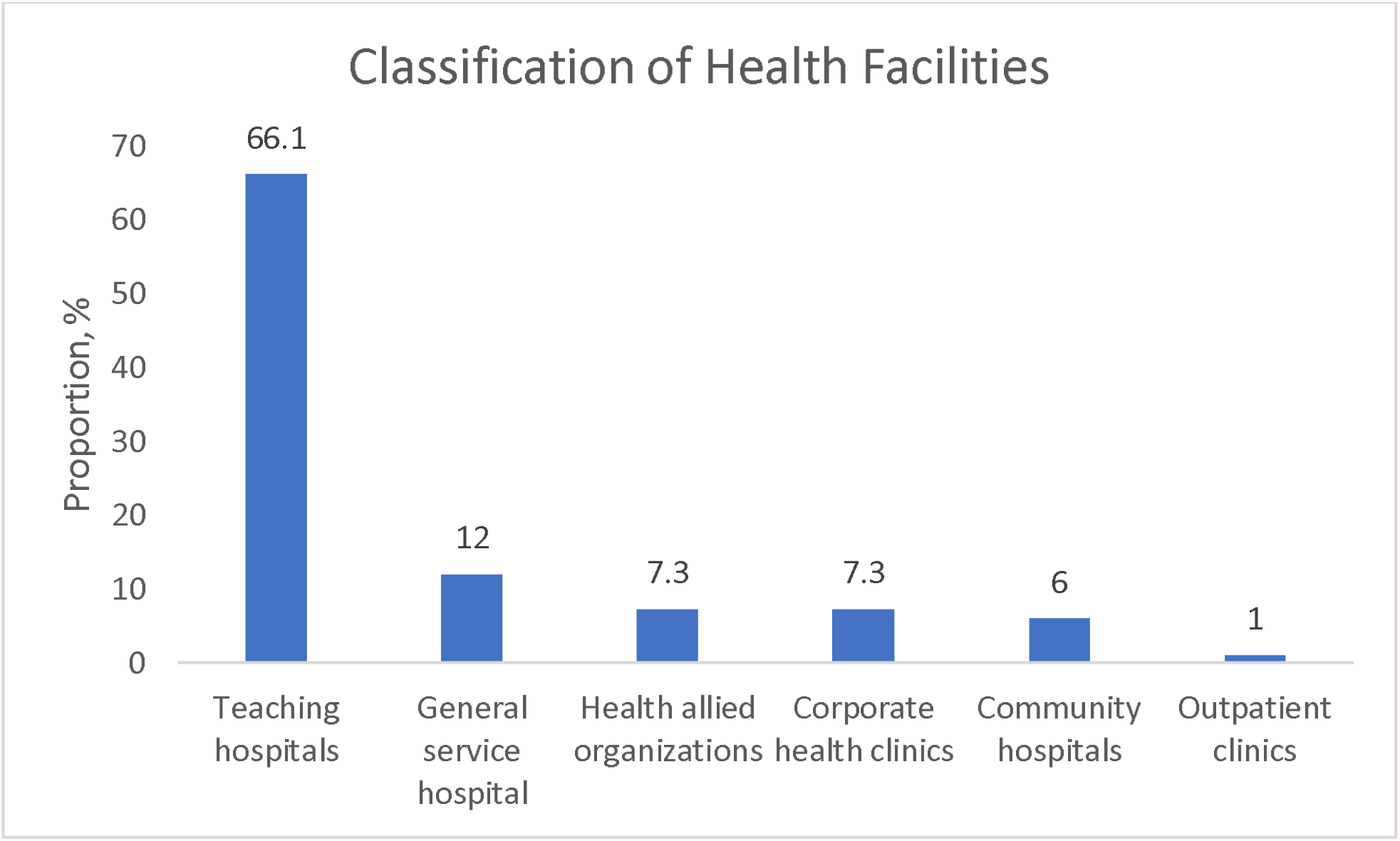
The proportion of infected healthcare workers by place of work.

### Predictors of illness severity and mortality amongst healthcare workers

The outcome proportion by risk factors is reported in Table 2. The effects of age, sex, and case class on illness severity were evaluated using both univariate and multivariate logistic regression (Table 4). Symptomatic cases were more likely to advance to severe illness (*χ*^2^(1) = 15.219, *α* = < 0.0001; aOR, 95% CI = 10.658, 2.494 – 45.552). The overall model was statistically significant (*χ*^2^(8) = 19.112, *α* < 0.0001) and explained 12.5% (Nagelkerke R^2^) of the variance in illness severity and correctly classified 89.4% of cases.

**Table 4.** Risk factors for COVID-19 illness severity among healthcare workers (n = 301)

| Variables | Univariate analysis | Multivariate analysis |
| --- | --- | --- |

| | $\chi^2$ | p-value | aOR | 95% CI | p-value |
| --- | --- | --- | --- | --- | --- |
| **Age** | 4.033 | 0.519 | 0.98 | 0.455 – 2.111 | 0.959 |
| **Sex<sup>a</sup>** | 0.004 | 0.952 | 1.003 | 0.971 – 1.036 | 0.859 |
| **Case class<sup>b</sup>** | 15.219 | <0.0001 | 10.658 | 2.494 – 45.552 | 0.001 |
| Classification table –89.4% correctly classified, constant = -4.139 |  |  |  |  |  |
| AOR – Adjusted Odds Ratio; 95% CI –95% Confidence intervals. |  |  |  |  |  |
| aRef group – female |  |  |  |  |  |
| bRef group – Asymptomatic |  |  |  |  |  |

Predictors of mortality assessed included age, sex, case class and illness severity (Table 5). The logistic regression model was statistically significant, *χ*^2,^(9) = 16.965, *α* = 0.049. The model explained 27.6% (Nagelkerke R^2^) of the variance in mortality and correctly classified 97.7% of cases. Age (*χ*^2^(5) = 13.7, *α* = 0.003; aOR, 95% CI = 1.079, 1.02–1.141 per year increase) was identified as a risk factor for mortality among healthcare workers with COVID-19 patients. The analyses on both illness severity and mortality are summarised in Table 3, 4, and 5, respectively. In both outcome analysis, age was categorised for univariate analysis.

**Table 5.**
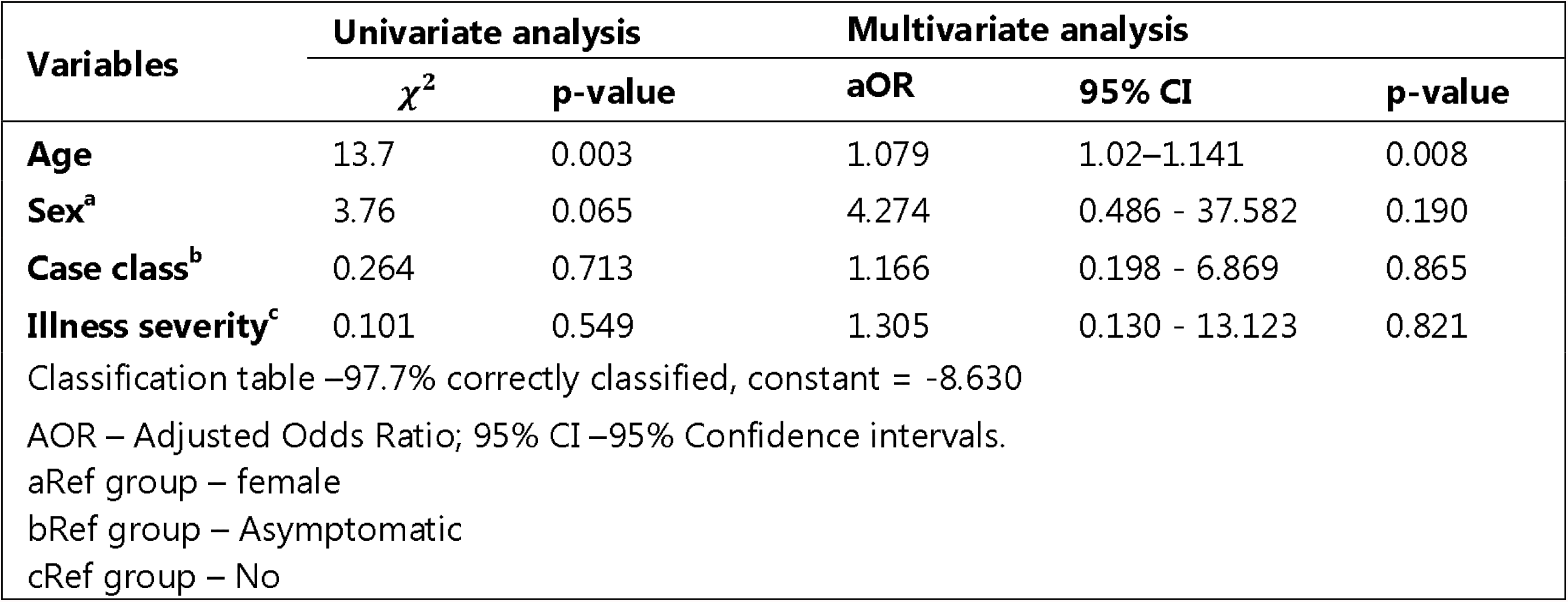
Risk factors for COVID-19 mortality amongst healthcare workers (n = 301)

## DISCUSSION

Using a comprehensive data on COVID-19 infections in healthcare workers in Rivers State Nigeria, this study showed a higher mortality of 2.3% in the study cohort compared to available evidence of 0.3%^10^. The difference in mortality is perhaps attributable to the geographical location of studies conducted. Studies on COVID-19 related mortality have mostly been conducted in developed countries (China, Italy, and USA), which showed lower mortality compared to the current study conducted in Nigeria, a developing country. Some of the known predictors of mortality amongst COVID-19 patients were also evaluated. COVID-19 infections that required hospitalisation was the measure of illness severity. Ten per cent of the study cohort experienced severe illness. The result agrees with available evidence from a meta-analysis that reported a 9.9% incidence of severe disease in healthcare workers ^10^.

In the evaluation of risk factors associated with COVID-19 severity and mortality, age and gender were significantly associated with COVID-19 mortality in healthcare workers. Age is a crucial risk factor in the epidemiology of COVID-19; prior research revealed patients above 65 years are at a greater risk of both disease severity and mortality from infection with COVID-19 ^11 12^. Consistent with research findings, mortality was higher in male patients in our study ^13 14^; although no significant association was deduced in the cohort evaluated. Also, infection amongst HCWs was typically asymptomatic with 89.4% not requiring hospitalisation; therefore, similar to conclusions from one study that observed less severe manifestations of COVID-19 infection in medical professionals ^15^. Our results showed significance between symptomatic cases and illness severity; however, more research is required to determine whether these findings are attributable to the healthy worker bias.

HCWs are the most important human resource for hospitals; the workplace-related mortality in HCWs not only compromise the workforce in healthcare settings but also affects the mental health of colleagues ^16 17^. A case fatality ratio (CFR) of 2.33% though comparable with global statistics for healthcare workers ^5^ is higher than both the CFR of the study area –Rivers state (0.98%) and Nigeria (1.23%) ^18^. There is a need for re-evaluation of compliance to the COVID-19 response protocol, the adequacy of personal protective equipment and working conditions in place for healthcare workers in Rivers state. Likewise, consideration must be given to the health-seeking behaviour of the healthcare workers in Nigeria and poor reporting of COVID-19 infection cases within this cohort. There is evidence that the practise of self-medication and reluctancy to obtain medical care is high among doctors and nurses in Nigeria ^19-23^. This behavourial pattern emphasises the need for more awareness and education on these issues within this group of healthcare professionals.

To our knowledge, this research is the foremost study representing a relatively comprehensive analysis of COVID-19 related mortality and disease severity in healthcare workers from available state records in Rivers state. Some of the limitations of this study include the reliance on reported infections and deaths, hence it’s impossible to estimate how many cases were missed by non-reporting. As an emerging research area in the current pandemic, there are other factors worth considering. For example, the effect of time of hospitalisation on disease severity and mortality. As a secondary analysis, we were unable to analyse this variable. Future studies to investigate this variable is essential.

## CONCLUSION

In conclusion, frontline healthcare workers are at an increased risk of exposure to COVID-19 infections. In Nigeria, there is the possibility of a higher risk of experiencing a severe disease if symptomatic while infected. It is imperative that preventive strategies, proper education, and awareness are put in place to protect healthcare workers.

## Data Availability

All data produced in the present study are available upon reasonable request to the Rivers State Ministry of Health

## ACKNOWLEDGEMENTS

We highly appreciate the editor and anonymous reviewers whose comments and suggestions helped greatly improve and clarify the manuscript. The authors acknowledge the Rivers State Public Health Emergency Operations Centre and the Rivers State Ministry of Health.

## CONFLICT OF INTEREST

The authors have declared no competing interest –financial or personal that could have influenced the work reported in this paper.

## FUNDING

The authors received no specific funding for this research.

## DATA AVAILABILITY STATEMENT

According to the Data Protection Law of the Federal Republic of Nigeria, the authors cannot publicly release the data accessed from the Rivers State Health Records Database. Data are available upon reasonable request with permission from the Rivers State Ministry of Health.

## Notes

### Funding Statement

This study did not receive any funding

### Author Declarations

Ethics Committee of the Rivers State Ministry of Health gave ethical approval for this work with Ethics ID: MH/PRS/391/VOL.2/809

